# Estimating number of cases and spread of Coronavirus disease 2019 (COVID-19) in the United Kingdom using critical care admissions, February to March 2020

**DOI:** 10.1101/2020.04.05.20054528

**Authors:** Mark Jit, Thibaut Jombart, Emily S Nightingale, Akira Endo, Sam Abbott, LSHTM Centre for Mathematical Modelling of Infectious Diseases COVID-19 Working Group, W John Edmunds

## Abstract

An exponential growth model was fitted to critical care admissions from multiple surveillance databases to determine likely COVID-19 case numbers and growth in the United Kingdom from 16 February – 23 March 2020, after which a national lockdown occurred. We estimate that on 23 March, there were 102,000 (median; 95% credible interval 54,000 - 155,000) new cases and 320 (211 - 412) new critical care reports, with 464,000 (266,000 – 628,000) cumulative cases since 16 February.

---

The first reported cases of Coronavirus disease 2019 (COVID-19) in the United Kingdom (UK) were identified in late January 2020 [1]. By 23 March, when a national lockdown prohibiting non-essential movement was announced, reported cases had increased to over 6000. However, reported cases are likely a small fraction of total infections because most COVID- 19 cases are mild or even asymptomatic [2]. Also, the proportion of infections that are reported changes by setting and over time, depending on the intensity of population-based surveillance, testing and contact tracing. However, COVID-19 patients admitted to critical care (high-dependency or intensive-care units, hereafter CC) offer a more stable indicator, since CC patients suspected of having COVID-19 have always been tested in the UK. We used two COVID-19 CC surveillance databases combined with data on reporting delays and disease severity to determine the likely number of and growth in infected people during the first phase of the COVID-19 epidemic in the UK. This work informed estimates produced by the Scientific Pandemic Influenza Group on Modelling (SPI-M) to advise the UK’s Department for Health and Social Care on its COVID-19 response.

## Data sources

Two data sources were used for CC case numbers:

i. The First Few Hundred (FF100) database containing virologically confirmed COVID-19 cases in the UK during the first phase of the epidemic. We extracted “sporadic” cases which were those identified through CC surveillance rather than contact tracing or targeted testing of travellers from high-risk countries, so that our analyses focussed on epidemic spread within the UK rather than including case importation. A few sporadic cases were identified through sentinel primary care testing rather than CC but could not be identified as such; however, there were only three such cases so numbers were too small to affect results. Sporadic cases were extracted from 16 February (the earliest date of onset for a sporadic case) to 6 March; cases peaked at 6/day on 6 March and then declined indicating that the FF100 was increasingly incomplete).
ii. CC cases in the COVID-19 Hospitalisations in England Surveillance System (CHESS) database which were collected on 15-20 March (the 7 cases reported on 13-14 March were ignored as the system was not fully operational then).

The age-dependent risk of an infected case being admitted to CC was estimated from fitting models to analyses of case data from China [3] and the USA [4] (details in Supplement S1).

## Modelling

The same model was fitted independently to incidence of patients admitted to CC based on their (1) date of symptom onset (for FF100 patients) or (2) date of admission (for CHESS patients, as date of symptom onset was not reported), using a Poisson likelihood with rate *Ae* ^*Bt*^, where *A* is the initial number of cases on 16 February and *B* is the growth rate of the epidemic. Delays between symptom onset and reporting were accounted for by fitting a Gamma distribution to time between symptom onset and CC admission for FF100 patients (see Supplement S2). When fitting modelled CC admissions to FF100, incidence was inflated by 18.6%compared to CC admissions fitted to CHESS, to adjust for the ratio of the population of the UK to that of England (the ratio of reported COVID-19 cases in the UK to that reported in England on 15-23 March would give a similar inflation factor of 19.8%).

*A* and *B* were sampled from their posterior distributions by using importance sampling; 10000 parameter sets for both were drawn from uniform distributions on [0,1] and then resampled with replacement at a probability for each sample weighted by the likelihood of that parameter set. Projected CC cases were then divided by age-dependent risks of CC admission to estimate actual infections in the population (Supplement S1).

## Results

We estimate that each COVID-19 case admitted to CC reported in FF100 and CHESS corresponds to a median of 89 (95% CrI 69 - 116) and 146 (95% CrI 90 - 274) infected individuals in the population respectively, based on Chinese and US severity data [3, 4]. Figure 1 shows the number of incident cases estimated on each day between 16 February and 23 March. On 23 March, 102,000 (95% CrI 54,000 - 155,000) new cases and 320 (95% CrI 211 - 412) CC reports are estimated to have occurred, with 464,000 (95% CrI 266,000 – 628,000) cumulative cases since 16 February. The best fitting exponential growth rates were consistent with an epidemic doubling time of 2.8 (95% CrI 2.4 – 3.1) days. Assuming an exponentially distributed serial interval of 4 days [5] gives an (approximate) reproduction number of 2.0 (95% CrI 1.9 - 2.1).

**Figure 1.**
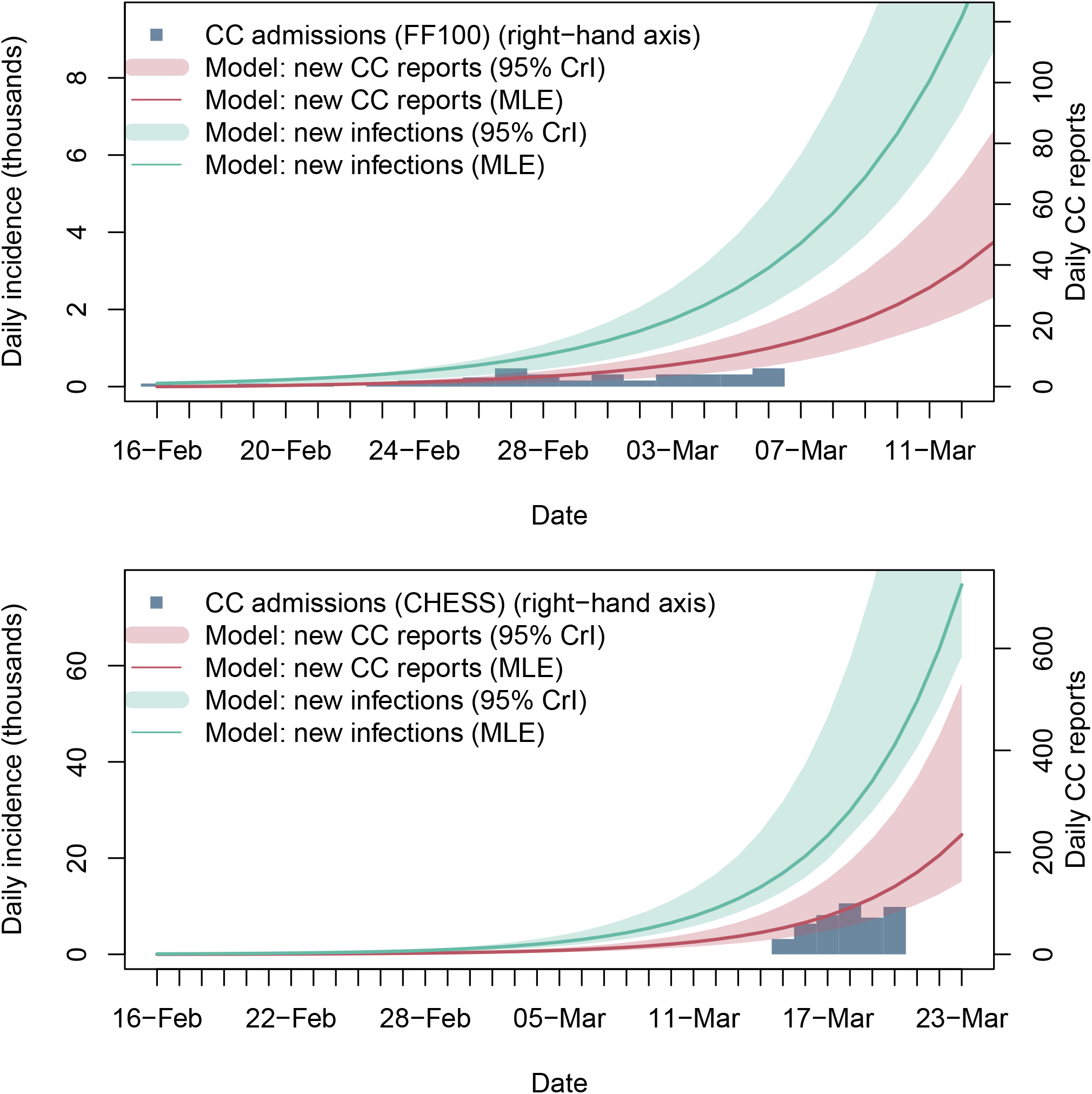
Model estimates of daily new COVID-19 infections and CC reports in the initial phase of the UK epidemic compared to data from FF100 (top) and CHESS (bottom, over longer time scale). The curves corresponding to the maximum likelihood estimate are in darker colours. Abbreviations: critical care (CC), COVID-19 Hospitalisations in England Surveillance System (CHESS). First Few Hundred (FF100), maximum likelihood estimate (MLE), 95% credible interval (95% CrI).]

Sensitivity analyses were performed on the datasets used for fitting, sensitivity of case detection and reporting, period of validity of the FF100 data and severity of COVID-19. Across the scenarios, the number of new infections on 23 March ranged from 92,000 (95% CrI 62,000 – 183,000) to 760,000 (95% CrI 753,000 – 1,020,000) (see Supplement S3).

For validation, we compared the median total number of CC admissions from 16-23 March (corresponding to an average duration of CC stay of 8 days in Wuhan data [6]) with the prevalence of COVID-19 patients in CC on 23 March (i.e. number of patients reported to be in CC) according to NHS England’s SitRep. Our model projects that 1261 (95% CrI 868 - 1500) patients were in CC on 23 March, about 75%higher than the figure of 714 from the Sitrep when inflated to account for the larger UK population.

## Discussion

Our results suggest that hundreds of thousands of COVID-19 infections had occurred in the UK by the time the national lockdown of 23 March was implemented, with incidence doubling every 2.8 (95% CrI 2.5 – 3.0) days. This suggests that only around 1%of infections were being detected and reported, since the official case count on 23 March was 6650 [7]. This provides evidence that strict physical distancing was necessary to prevent health services from being overwhelmed. However, across all scenarios the majority of the UK population remained uninfected, and hence timely interventions to reduce physical contact could have a large impact. Growth of the COVID-19 epidemic beyond 23 March depends on the effectiveness of these interventions.

Our investigations revealed weaknesses in two early sources of CC surveillance (FF100 and CHESS). In particular, the number of cases reported in the two databases markedly differ, with the FF100 reporting far fewer CC cases (e.g. 6 cases on 6 March from FF100 compared to 100 on 18 March from CHESS) even after accounting for different periods of coverage and delays to reporting. Further analysis also suggested delays between the date of being reported as a case and the date on which a case actually appears in the FF100 database which made correcting for right-truncation of cases difficult (see Supplement S2). Also, the proportion of cases estimated to require CC treatment is substantially lower than observed elsewhere (and hence the multipliers are higher) because of the young profile of CC cases in both FF100 and CHESS. For example, 34%of CC patients were below 45 years old in CHESS compared to only 12%in published US data up to March 16 [4]. This may reflect biases in CHESS reporting, differential propensity to test by age, a higher triage threshold for CC admission of older patients than in the USA, or be an indication of hospital-based COVID-19 transmission affecting mainly younger children.

However, the overall conclusion that COVID-19 daily incidence in the UK on 23 March was in the hundreds of thousands of infections and cumulative incidence approaching a million appears to be robust even when challenged by a range of sensitivity analyses to account for different assumptions. Furthermore, the order of magnitude is on the same order of magnitude as data from SitReps that have less granularity but possibly better coverage. Lower numbers in the SitReps may reflect decreasing propensity to admit patients as CC beds become filled. Our investigations highlight the usefulness of CC surveillance in understanding epidemic dynamics and informing response measures, and hence the need for timely and complete reporting over the course of the epidemic.

## Data Availability

All data generated by the model is shown in the tables and figures; detailed numerical results are available from the authors on request. Data inputs to the model should be obtained by contacting the custodians of the data.

https://github.com/markjit1/covid19_nowcast

## Acknowledgments

This research was funded by the National Institute for Health Research - Health Protection Research Unit in Immunisation (MJ) and for Modelling Methodology (MJ, TJ, JE), the Economic and Social Research Council RCUK grant ES/P010873/1 (TJ), the UK Public Health Rapid Support Team (TJ), the Bill & Melinda Gates Foundation grant OPP1183986 (ESN) the Nakajima Foundation (AE), the Alan Turing Institute (AE) and the Wellcome Trust grant 210758/Z/18/Z (SA). The views expressed in this publication are those of the author(s) and not necessarily those of the NIHR, Public Health England or the UK Department of Health and Social Care.

We would like to acknowledge the other members of the London School of Hygiene & Tropical Medicine COVID-19 modelling group, who contributed to this work. Their funding sources are as follows: Hamish Gibbs (NIHR ITCRZ 03010), Kathleen O’Reilly (BMGF OPP1191821), Joel Hellewell (Wellcome Trust 210758/Z/18/Z), Alicia Rosello (NIHR PR-OD- 1017-20002), Billy Quilty (NIHR 16/137/109), Charlie Diamond (NIHR 16/137/109), Petra Klepac (BMGF INV-003174), Amy Gimma (RCUK/ESRC ES/P010873/1), Rosalind M Eggo (HDR UK MR/S003975/1), Megan Auzenbergs (BMGF OPP1191821), Samuel Clifford (Wellcome Trust 208812/Z/17/Z), Gwen Knight (MRC MR/P014658/1), Sebastian Funk (Wellcome Trust 210758/Z/18/Z), Fiona Sun (NIHR 16/137/109), Jon C Emery (ERC #757699), Kiesha Prem (BMGF INV-003174), Yang Liu (BMGF INV-003174, NIHR 16/137/109), Kevin van Zandvoort (R2HC), Christopher I Jarvis (GCRF ES/P010873/1), James D Munday (Wellcome Trust 210758/Z/18/Z), Adam J Kucharski (Wellcome Trust 206250/Z/17/Z), Carl A B Pearson Gates (BMGF OPP1184344), Timothy W Russell (Wellcome Trust 206250/Z/17/Z), Nikos I Bosse (Wellcome Trust 210758/Z/18/Z), Stefan Flasche (Wellcome Trust 208812/Z/17/Z), Rein M G J Houben (ERC #757699), Simon R Procter (BMGF OPP1180644), Nicholas Davies (NIHR HPRU-2012-10096).

## Authors’ contributions

Conceptualisation: MJ, JE; model formulation: MJ, JE, TJ, ESN, AE, SA; analysis and code development: MJ, TJ, ESN, AE, SA; writing: MJ, JE, TJ, ESN, AE, SA. The members of the Working Group contributed equally in model formulation, analysis, writing reviewing the manuscript and approving the work for publication. The order was assigned randomly.

## Code availability

Computer code used for the analysis is available on https://github.com/markjit1/covid19_nowcast.

